# Emergency medicine patient wait time multivariable prediction models: a multicentre derivation and validation study

**DOI:** 10.1101/2021.03.19.21253921

**Authors:** Katie Walker, Jirayus Jiarpakdee, Anne Loupis, Chakkrit Tantithamthavorn, Keith Joe, Michael Ben-Meir, Hamed Akhlaghi, Jennie Hutton, Wei Wang, Michael Stephenson, Gabriel Blecher, Paul Buntine, Amy Sweeny, Burak Turhan, On behalf of the Australasian College for Emergency Medicine, Clinical Trials Network

## Abstract

**Objective:** Patients, families and community members would like emergency department wait time visibility. This would improve patient journeys through emergency medicine. The study objective was to derive, internally and externally validate machine learning models to predict emergency patient wait times that are applicable to a wide variety of emergency departments.

**Methods:** Twelve emergency departments provided three years of retrospective administrative data from Australia (2017-19). Descriptive and exploratory analyses were undertaken on the datasets. Statistical and machine learning models were developed to predict wait times at each site and were internally and externally validated. Model performance was tested on COVID-19 period data (January to June 2020).

**Results:** There were 1,930,609 patient episodes analysed and median site wait times varied from 24 to 54 minutes. Individual site model prediction median absolute errors varied from +/−22.6 minutes (95%CI 22.4,22.9) to +/− 44.0 minutes (95%CI 43.4,44.4). Global model prediction median absolute errors varied from +/−33.9 minutes (95%CI 33.4, 34.0) to +/−43.8 minutes (95%CI 43.7, 43.9). Random forest and linear regression models performed the best, rolling average models under-estimated wait times. Important variables were triage category, last-k patient average wait time, and arrival time. Wait time prediction models are not transferable across hospitals. Models performed well during the COVID-19 lockdown period.

**Conclusions:** Electronic emergency demographic and flow information can be used to approximate emergency patient wait times. A general model is less accurate if applied without site specific factors.

**What is already known on this subject:**

⍰ Patients and families want to know approximate emergency wait times, which will improve their ability to manage their logistical, physical and emotional needs whilst waiting
⍰ There are a few small studies from a limited number of jurisdictions, reporting model methods, important predictor variables and accuracy of derived models

**What this study adds:**

⍰ Our study demonstrates that predicting wait times from simple, readily available data is complex and provides estimates that aren’t as accurate as patients would like, however rough estimates may still be better than no information
⍰ We present the most influential variables regarding wait times and advise against using rolling average models, preferring random forest or linear regression techniques
⍰ Emergency medicine machine learning models may be less generalisable to other sites than we hope for when we read manuscripts or buy commercial off-the-shelf models or algorithms. Models developed for one site lose accuracy at another site and global models built for whole systems may need customisation to each individual site. This may apply to data science clinical decision instruments as well as operational machine learning models.

## Main Manuscript

### Introduction

Deciding where to seek care for acute medical problems is complex and nuanced. Many decisions are made with limited information and at times little transparency from health services. Most people hope to be seen by a definitive provider immediately on arrival but usually have to wait for treatment. Emergency department (ED) proximity and wait times are the major influencers on patient choice of facility.[1–4] Wait time visibility assists with meeting physical, logistic and psychological needs of patients.[5] There is increasing consumer advocacy for transparency of information about health service resources. There is also health service interest in displaying wait times. Many examples exist in the USA, Canada and emerging interest has been seen in Australia and other jurisdictions.

Information technology (IT) capabilities and applied data science techniques are becoming increasingly available to acute care services. Many emergency departments collect a large volume of electronic point-of-care patient data, relating to demographics, flow and clinical care. In a community where data from multiple emergency departments are available, knowledge of queue lengths could facilitate optimal patient load-balancing across acute care facilities. This has the potential to reduce the harms of long waits.

Previously published information is available regarding how to predict wait times in emergency medicine. Manuscripts report a variety of predictor variables, model techniques and accuracy, from either a single centre or small number of sites. Sun et al[6] used quantile regression techniques in a single emergency department, and found that by using triage categories, the number of unseen patients and the number of new patients treated by physicians in the last hour, they could predict wait times to an accuracy of +/− 12 minutes.

Ang et al[7] compared statistical to machine learning models using data from four emergency departments in the USA in response to inaccurate commercial wait time predictors, finding Q Lasso to be the best model with less under-estimation of wait times; using only time-of-day and day-of-week data. Arha[8] used a tree-based regression model, with simple predictor variables available at triage. Senderovich et al[9] found that by adding congestion variables, patient flow predictors had increased accuracy.

There is limited knowledge regarding wait time predictors performance across a variety of jurisdictions, patient catchments and healthcare resources. There is no knowledge about whether one model might be able to be applied across multiple emergency departments for system-wide implementation or how predictive models perform during unexpected events with variations in demand (e.g., COVID-19).

### Objectives

The primary objective of the study was to develop and internally validate predictive algorithms for patient wait times (triage-to-provider). Secondary objectives include determining the relative importance of each predictor variable and model method, whether models are transferable across different emergency departments (external validation), and the performance of the models during special events (COVID-19).

## Methods

### Study design and Setting

This is an observational study using retrospective administrative data to develop, compare, and validate prediction models for patient wait times at emergency departments. Data from 2017 to 2019 from 12 emergency departments were used for the main study, followed by data from January to June 2020 from three emergency departments to test performance during COVID-19 conditions.

Mandatory point-of-care emergency patient demographic, flow and clinical data are collected for every patient by clerks and clinicians in Australia. In Victoria, these defined data populate the governmental Victorian Emergency Minimum Dataset (VEMD).[10] Data available at time of triage were used as predictor variables.

There are 24 million residents in Australia and emergency medicine manages eight million patient episodes annually. The majority (93%) of Australian residents attend government-funded, public emergency departments with no patient co-payments. There are no restrictions on individual choice of emergency department. Regional to tertiary emergency departments were invited to participate if they were part of an academic health science centre or were engaged via research networks. Ten Melbourne and two Queensland emergency departments participated, comprising one private, one paediatric, four major, two large metropolitan, and four medium metropolitan hospitals. Hospital #7 (H7) displayed predicted patient wait times (prior in-house models) online, in their waiting room, and to Ambulance Victoria during the study.

The study received Monash Health ethics committee approval (RES-19-0000-763A).

### Data sources and Measurements

Electronic medical record software applies time-date stamps to clinician activities (e.g., triage). Clerical staff collect demographic data from patients at initial registration. Clinical staff record data whilst attending to a patient. The VEMD datasets from each hospital were the primary source of data for this study. VEMD data are routinely checked for completeness, accuracy and administrative errors by an emergency physician at each site prior to submission to the Victorian Government.

Three years of retrospective, de-identified VEMD data were obtained from twelve hospitals in Australia (mainly Melbourne). Hospital names were replaced with alphanumeric codes prior to analyses. All episodes of care were eligible for inclusion in the study. Data were collected in early 2020 and arranged into training (2017 and 2018) and testing datasets (2019), maintaining the temporal order based on patient arrival times. The training dataset was used for exploratory analysis and learning prediction models. The testing dataset was used to internally and externally validate the prediction models. Further retrospective data were collected from three emergency departments, for the period of January 1st 2020 to June 30th 2020 coinciding with major variations in emergency attendances secondary to COVID-19 concerns (April, May 2020). These data were used to evaluate the stability of model performance during unexpected circumstances.

### Variables

Variables used in this study are presented in Appendix 1a. Variables collected after triage/registration were excluded from the models, except those required for calculation of the dependent variables. We used 19 predictor variables (13 VEMD and six derived) in total.

### Outcomes

The primary outcomes of this study were triage-to-provider wait times for all patients, predicted at triage. Secondary outcomes included the accuracy of each predictive model (internal validation); determining if a global model or individual models performed better; identifying the best technique to generate these models; the relative contribution of each variable to the models; assessment of how each model performs at different sites (cross-site, external validation) and evaluation of how the models perform during COVID-19 conditions or unusual circumstances. Researchers weren’t blinded to outcomes. The outcome choices and definitions were informed by a large, multisite, qualitative study of community members, consumers, paramedics and health administrators.[5] These participants recommended a prediction accuracy of +/− 30 minutes (unpublished data).

### Analysis

#### Study size

We used time-based sampling similar to previous studies of wait time prediction that have used time periods ranging from one month to one year.[6–8] We obtained three years of data from each hospital to account for seasonal variations in patient visits. Multiple hospitals were enrolled to allow cross-site validation evaluations. The accepted convention of using a minimum of 30 to 50 data points per variable was applied.

#### Data cleaning, outliers and missing data

Patient data rows were checked for missing values related to the primary outcomes and episodes were removed from analysis if the primary outcome variables were missing. We therefore removed patients who left without being seen by a provider (n=133,204 (6.85%)). Other missing values were replaced with “unknown” or “other” categories using VEMD descriptors. Three hospitals did not collect ambulance data. The total number of unique patient episodes where the value of at least one of the predictor variables is “unknown/other” is (n=1,733,247), covering a total of eight predictor variables (Appendix 1b).

Negative values for triage-to-provider time (n = 236 (0.01%)) were removed from the analysis. We also removed patient data where the wait time exceeded the maximum of 360 minutes and the predefined statistical outlier threshold value (defined as 1.5 times the interquartile range (IQR = Q3 - Q1) over Q3) which were mainly generated by administrative data entry errors for triage-to-provider time (n = 13,612 (0.7%)).

#### Standardising and encoding data

Hospitals providing non-VEMD formatted data (Queensland) had their data converted to VEMD format. One-hot encoding[11] was applied to all categorical variables prior to prediction model development as all categorical variables were nominal with the exception of triage category. We assessed that it was preferable to lose order information for triage by applying one-hot encoding than to treat triage category as a continuous variable as the distances within levels of triage category were non-linear.

#### Model building and recalibration

Python scikit-learn and statsmodels modules were used for machine learning model development. Guided by wait-time prediction literature,[7–9,12] we used three statistical and machine learning techniques (i.e., linear regression, random forests, and elastic net regression) and a rolling average approach (i.e., the average calculation of the outcome of previous k = 4 observations). We included all predictor variables in model construction and undertook a post-hoc variable importance analysis. We rebuilt the models with the most important variables only and compared the performance of the simplified models to the initial models. To foster future replications, we provide code snippets for model construction in an online repository: https://doi.org/10.5281/zenodo.459978.

The “last-k” variable is the average triage-to-doctor wait time for the last “k” patients seen by a provider. To determine the appropriate value of “k” for this study, we performed a sensitivity analysis by observing the performance of prediction models constructed using different k values (i.e., 3-10). We found that the performance differences are statistically indistinguishable across different k values. We selected the k value of 4 for this study, which produced models with the best performance.

#### Validation

For site-specific accuracy testing (internal validation), we used a time-wise hold-out validation approach.[13] Patient records were sorted for each hospital by their arrival time. Data from 2017-18 were used to construct site-specific prediction models for all individual hospitals, while 2019 data were used to evaluate prediction models within each hospital. For cross-site comparisons of site-specific models, we used a time-wise cross-site approach.[19] The site-specific models were tested using 2019 data from other hospitals (e.g., train with Hospital A data from 2017-18, then test with Hospital B data from 2019), resulting in 132 pairwise combinations.

We undertook geographical cross-state testing (external validation). Global prediction models were constructed using 2017 and 2018 data from hospitals in Victoria and evaluated using 2019 data from Queensland. We also tested the global model performance against combined 2019 Victorian all-site data.

We used two boosting ensemble techniques (light GBM and eXtreme gradient boosting) and one hyperparameter optimised implementation of random forests (Random Forests (HPO)) in our model validation. We found non-statistically significant improvements using these tools and as they came at considerable computational cost, we excluded them from the main analysis.

For validation during unexpected events, we compared model accuracies between the first six months of 2019 (surrogate for normal conditions) and the first six months of 2020 (surrogate for unexpected events, e.g. COVID-19), using data and models from three emergency departments.

To assess model performance, we calculated the Absolute Errors (AE) between the actual time and the predicted time for all models and hospitals. We then calculated the median of these distributions of Absolute Errors (MAE) to identify the best model for and across 12 hospitals.

### Statistical methods

Scott-Knott effect size difference test was used to identify performance ranking, based on MAE, of the prediction models for internal validation.[15] The Scott-Knott effect size difference test is a multiple comparison approach that produces statistically distinct and non-negligible (effect size) groups of distributions. We used the implementation provided by the sk_esd function of the ScottKnottESD R package version 2.0.3. Mann Whitney U tests were used to identify whether the performance (MAE) difference between two models was statistically significant, then Cliff’s delta tests,[16] ldl, were used to measure the effect size. The interpretation of Cliff’s delta values is as follows: ldl < 0.147 negligible, ldl < 0.33 small, ldl < 0.474, otherwise large.[17] We used cliff.delta function of the effsize R package version 0.7.8 for calculating Cliff’s delta. For all statistical tests, we used a statistical significance level of □ = 0.05 and sought non-negligible effect sizes.

### Patient and public involvement

The primary outcome of this study was determined by a qualitative study involving patients, the public and other stakeholders.[5] Consumers and community stakeholders contributed to the design and write up of the study.

## Results

### Characteristics of study subjects

Twelve emergency departments contributed data. Two sites were unable to obtain ethics approval (regional referral, medium metropolitan). Flow through the study is presented in Figure 1. Department and patient demographics are presented in Table 1. The total number of patient episodes included in the study were 1,930,609 with 1,388,509 in the training and 542,100 in the testing datasets. Overall admission rates were 29% and 23% of patients arrived by ambulance.

**Table 1.**
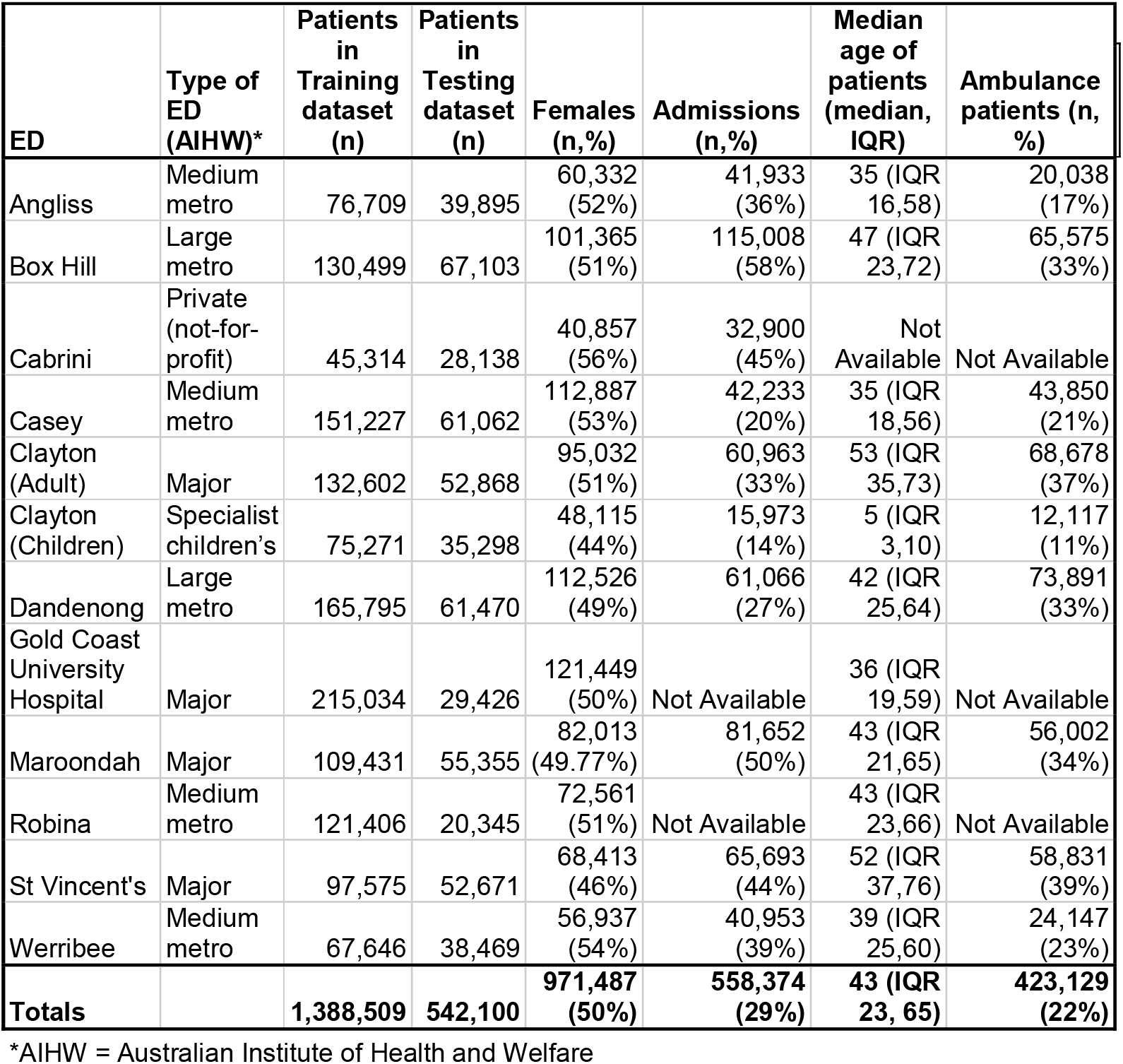
Emergency department and patient demographics

**Figure 1.**
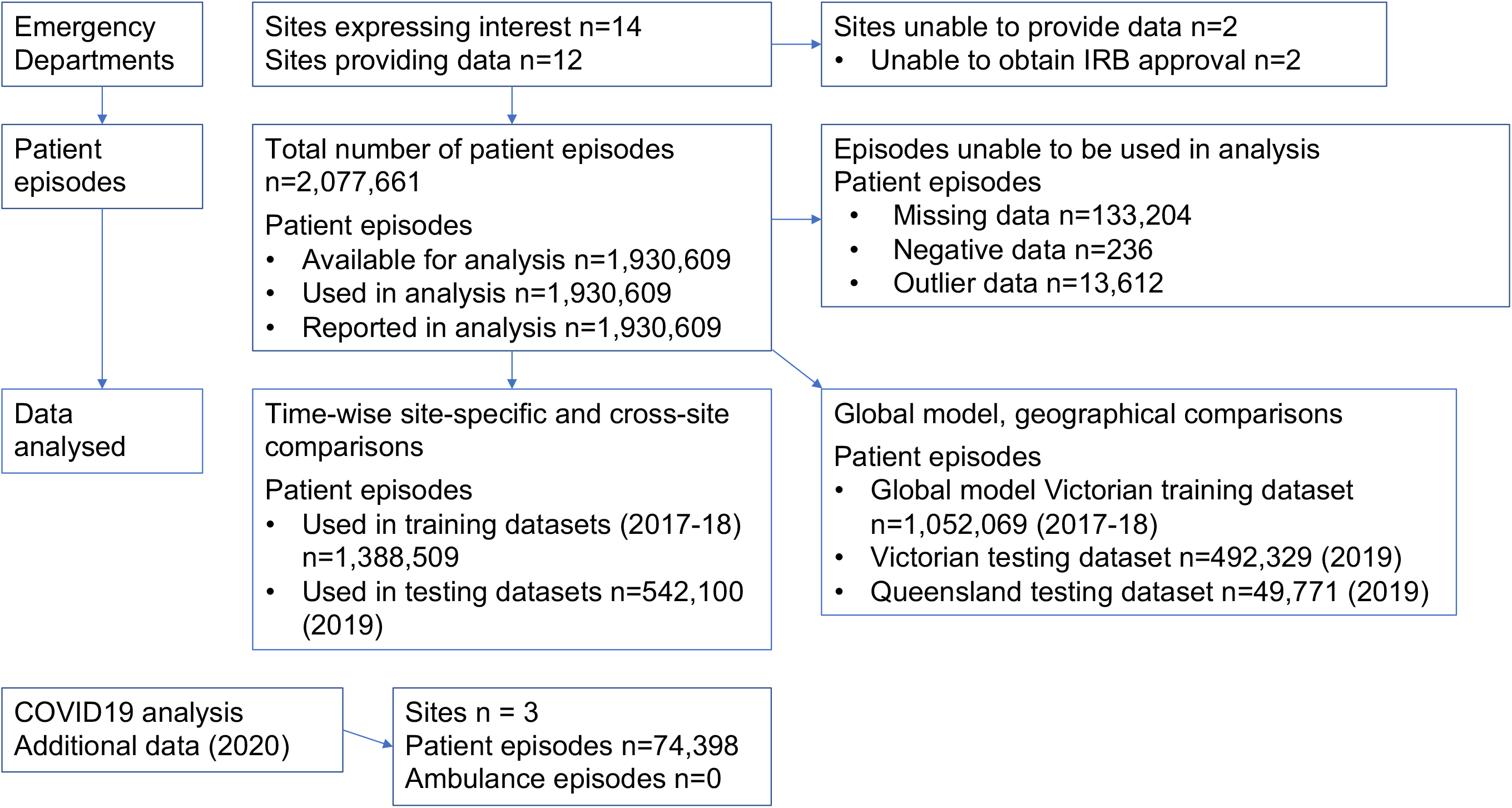
Participant flow through study

Wait time proportional distributions were similar throughout sites, although specific wait times at each site varied, with a median site range of 24-54 minutes for triage-to-provider time (Figure 2). Distribution of outcomes are right skewed but we did not apply any transformation to require positive predictions, since predictions of negative values are expected to be rare events and can be replaced with zero in deployment.[6]

**Figure 2.**
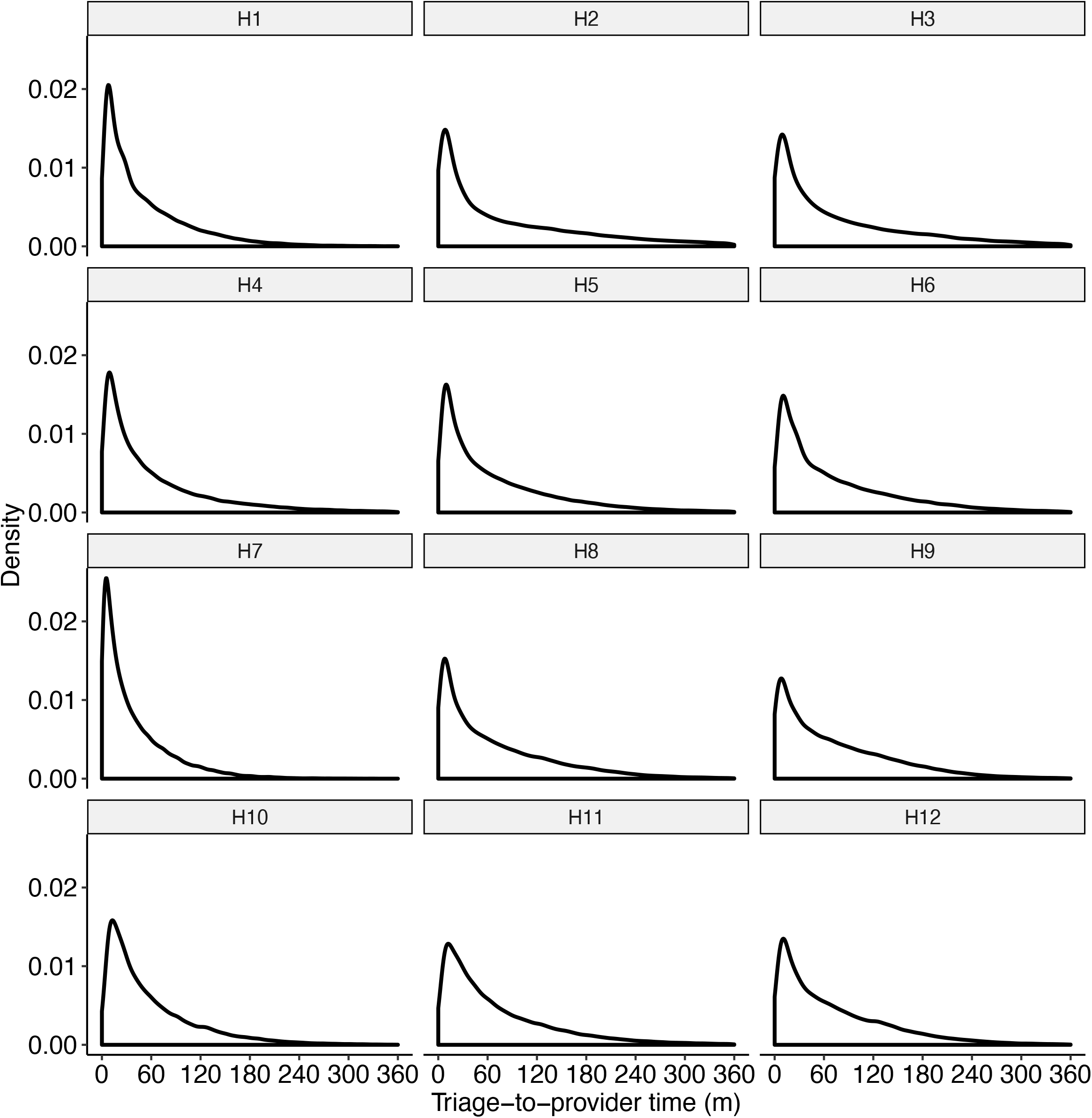
Site distribution of triage−to−provider wait times.

## Main results

### Internal validation of site-specific models (same hospital, using later time period)

The performance rankings produced by the Scott-Knott effect size difference test showed that Random Forests and Linear Regression performed the best (1^st^ rank) for all studied hospitals (n=12) followed by Elastic Net (n=11) and Rolling Average (n = 5). Random Forests, Linear Regression, and Elastic Net outperformed Rolling Average at seven hospitals. The Median Absolute Error (MAE) of Random Forests varied from 22.6 minutes (95% CI = [22.4, 22.9]) for H7 to 44.0 minutes (95% CI = [43.4, 44.4]) for H2. The distributions of the absolute errors of internal validation for ED wait-time (triage-to-provider) prediction are shown (Figure 3). The prediction models predicted a wait time to within +/− 30 minutes of the actual wait time between 40 and 63% of the time, depending on the site.

**Figure 3.**
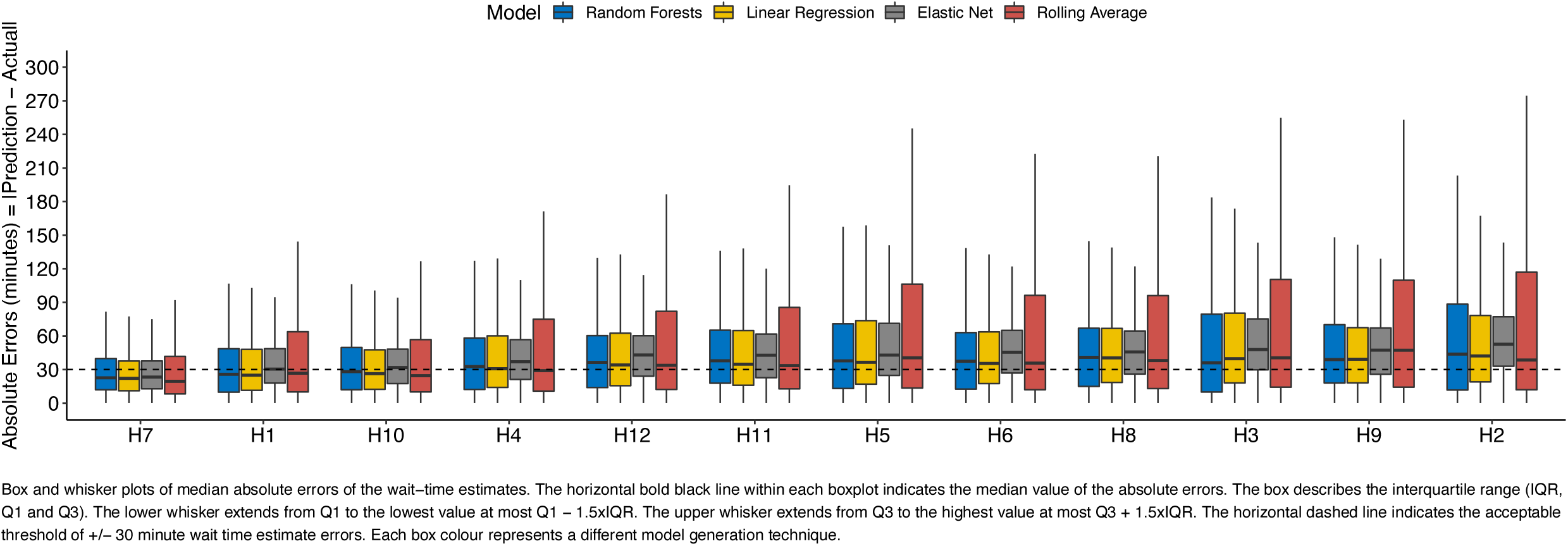
Distributions of the Absolute Errors (AE) for triage−to−provider time (Internal Validation, site−specific models).

The performance differences between Random Forests and Linear Regression were negligible for all hospitals according to Cliff’s delta effect size. Rolling average models consistently underestimated patient wait times. More details regarding the actual errors are shown in Appendix 1c.

### Variable Importance Analysis

Triage Category (median importance score = 65%, IQR (54%, 74%)), Arrival Time (Hour) (median importance score = 15%, IQR (12%,25%)), and the average wait time of the last k-patients (median importance score = 15%, IQR (7%,21%)) were the three most important Random Forest predictive variables across all sites. The distributions of importance scores are shown in Figure 4.

**Figure 4.**
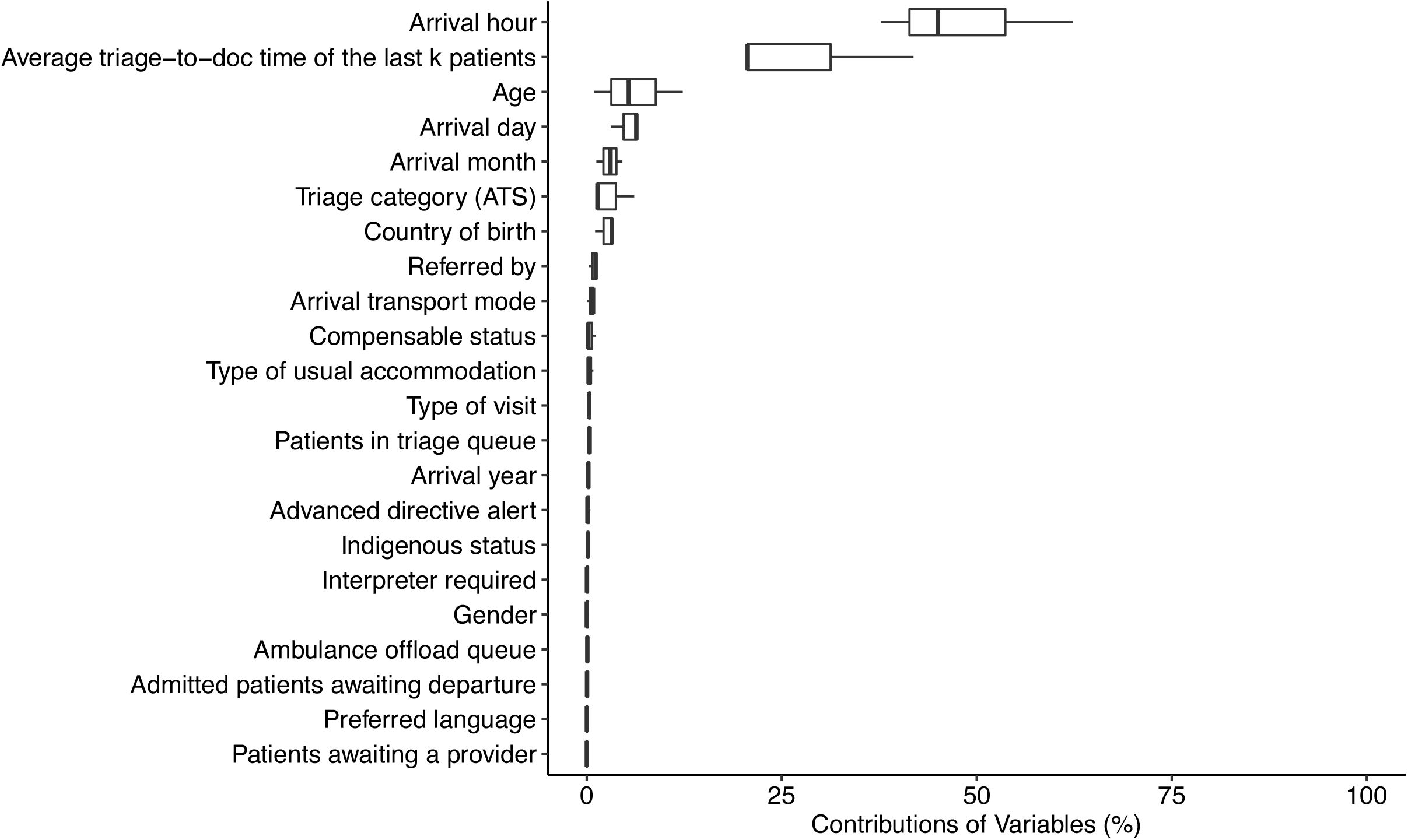
Distributions of importance scores for each variable.

Simplified models were built with the top-ranked variables, which accounted for 95% of the relative variable importance. They demonstrated similar accuracy to full models with all variables. Performance differences between Random Forest simplified models and full models were not statistically significant and had negligible effect sizes for all hospitals. Distributions of the absolute errors of these simplified models are shown in Appendix 1d.

### Site-specific model performances at other single sites

Single site models perform better for the specific hospital they were developed for, compared to other sites. Out of 132 pairwise Random Forest combinations, 119 combinations had statistically significant performance differences with negligible to medium effect size. Ninety-seven (~82%) yielded higher errors by 0.02-39.6 minutes and 22 (~20%) yielded lower errors by 0.01-12.4 minutes compared to their site of origin. This suggests that site-specific patient wait time prediction models are not transferable to different hospitals. The distributions of absolute errors for triage-to-provider time prediction are shown in Appendix 1e.

### Global model performance

Multi-hospital global models that were constructed from Victorian 2017-18 data performed similarly when tested with 2019 data from hospitals in Victoria and Queensland. The global models didn’t perform as well as site-specific models. The Median Absolute Errors (MAE) of these global models varying from 32.2 minutes (95% CI = (32.1, 32.3)) for Linear Regression to 42.6 minutes (95% CI = (42.5, 42.7)) for Elastic Net when tested with Victorian hospital data, and varying from 36.1 minutes (95% CI = (35.6, 36.6)) for Linear Regression to 42.4 minutes (95% CI = (42.1, 42.7)) for Elastic Net when tested with patients from hospitals in Queensland. The distributions of the absolute errors of these global models are shown in Figure 5.

**Figure 5.**
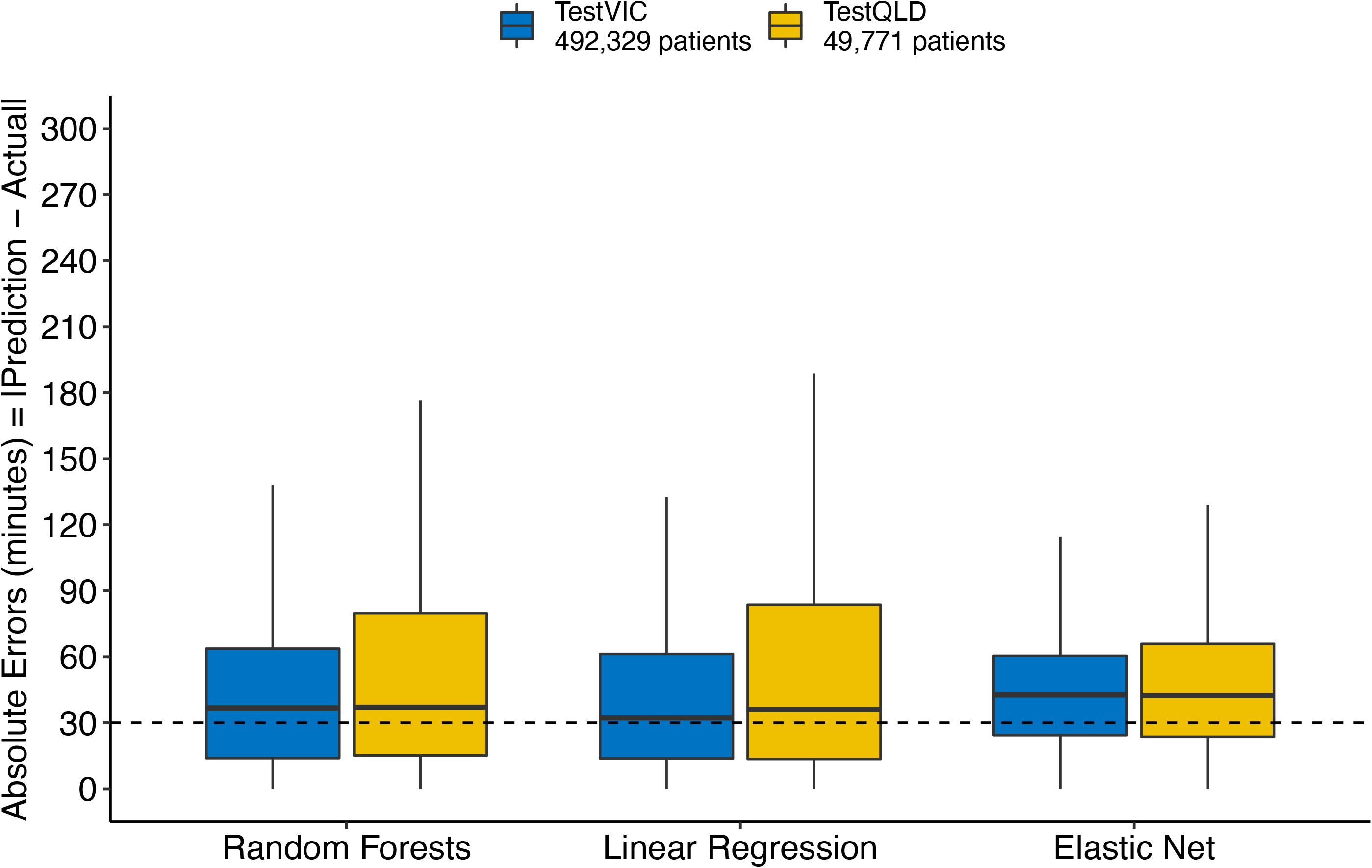
Distributions of Absolute Errors (AE) of global models.

### Impact of COVID-19 on model accuracy

Three hospitals provided 2020 data (n = 74,398) covering some of the reduced attendances COVID-19 period in Victoria. We observed that patient wait time models that were built using past data from 2017 and 2018 still performed at reasonable accuracy with MAE differences ranging from 0.03-6.0 minutes. Though these performance differences are statistically significant, except for Random Forests at H10 and Elastic Net at H12, the effect sizes were all negligible for these hospitals. Three models yielded higher errors by 1.7-5.00 minutes and four yielded lower errors by 0.5-6.0 minutes. Distributions of the absolute errors of models during COVID-19 are shown in Appendix 1f.

## Limitations

Limitations of the study include using only administrative demographic and ED flow data, and using only Australian data. There are no direct measures of resource availability or processes within the ED (e.g. nursed cubicles, streaming within the ED); hospital capacity (e.g. available beds) or community resourcing (e.g. ambulances to transport patients to nursing homes). There were also no measurements of patient comorbidity or diagnosis used in the models. Inclusion of this information may improve prediction accuracy in future models. Excluding did not wait patients or including triage category 1 patients from the study may have over or underestimated true wait times. Additionally, we don’t have information about how this model would perform during a disaster with a rapid surge in attendances. Models present general estimates of patient wait times for those arriving at the emergency department, they don’t generate individualised wait times for each patient.

Models can generate nonsensical outputs, for example linear regression can generate negative predictions which do not make practical sense. In practice, negative predictions can be replaced with 0. We observe the best prediction results in H7 which is the only hospital where wait-time predictions, from an in-house developed model, are shared on their website and on site. This may have affected the behaviour of lower acuity patients in choosing to visit H7 with more flexibility, at times when less waiting is expected, which could have resulted in a more homogenous – and easier to model – distribution than the other hospitals in our sample. Machine learning models may or may not introduce or amplify bias in healthcare. There are currently no reliable ways of testing for bias in machine learning when applied to healthcare datasets (personal communication, Dr Aldeida Aleti, Monash University) and so we are unable to determine if these outputs are biased for or against any particular group of patients.

## Discussion

In summary, using emergency patient demographic and flow data from 12 studied hospitals, it is possible to build triage-to-provider predictive waiting time models to an accuracy of +/− 22.6 to 44.0 minutes. Predictions were within +/− 30 minutes of the actual wait time between 40% and 63% of the time, varying by site. The best performing models used Random Forests and linear regression methods for triage-to-provider prediction. Average wait time of the last k-patients, triage category and patient arrival time were the most important predictor variables. Accuracy is reduced when a model developed for one site is used at another site or a global model (developed by multiple sites) is used. When special events occur such as COVID19 reduction in attendances, prediction accuracy is maintained.

To our knowledge, this is the largest study of its kind to date. The accuracies obtained for triage-to-provider times (23 to 44 minutes) are less than those reported from a Singapore study.[6] Sun et al. built different models for each acuity level and this approach to prediction at a finer granularity level may explain the performance difference. They omitted acuity category 1 and reported accuracies of 11.9 minutes for acuity category 2 and 15.7 minutes for acuity category 3. We chose a single prediction based on consumer feedback that triage categories are not understood by patients and families.[5] Ang et al. modelled low-acuity patients only and reported performance in terms of Mean Squared Error, arguing that median based measures tend to underestimate due to right skewed distribution of wait-times.[7] We observed this with Rolling Average models, but not others. Ang et al. reported 9.4 minutes for non-absolute median error, which we outperform at a range of 1.1 to 8.9 minutes depending on the site, even after inclusion of all triage categories in the analyses.[7]

Importantly, we found that using a model developed for one site will come at a cost in accuracy for the new site, prompting caution prior to promoting a “one size fits all” model. Models developed for one site can be applied at other sites, but lose accuracy. The global models may reduce time and cost spent developing individual models but are less accurate at individual sites. External validation of data science models should be undertaken prior to implementation at new jurisdictions, particularly where clinical care decisions might be assisted by machine learning algorithms or models.

This is the first literature describing how models perform during unusual events. We demonstrated that COVID19 lockdowns didn’t have a negative impact on model accuracy. In Australia, this period of time was one of both significantly reduced emergency attendances and reduced productivity for physicians due to the increased complexity of managing patients and departments.[18] These data don’t cover periods of surge.

Qualitative work has shown that patients want access to wait times and would use times to address a large variety of needs.[5] This study has shown that it is possible to predict approximate wait times, however it has also demonstrated that the range of predictions is less accurate than desired by stakeholders. Some information about wait times may still be useful to patients, even if the prediction range is broad. Overestimated and underestimated predictions may be perceived differently by patients and families. Overestimated predictions may be perceived positively if patients wait a shorter time than predicted, but could deter patients from seeking care. Random Forests tend to overestimate more than linear regression. Rolling average models underestimated wait times the most (Figure 5) and could either make emergency flow seem better than it is, or generate dissatisfaction from patients when waits exceed predictions.[19–23]

In summary, using limited data available at point-of-care, wait times can be predicted to +/− 23 to 44 minutes. Models should be individually built for each hospital and are likely to perform the same during COVID-19 like conditions.

## Data Availability

Data are not available for sharing.
To foster future replications, we provide code snippets for model construction in an online repository.

https://doi.org/10.5281/zenodo.459978

## Author contributions

Principal Investigator: KW; Funding: KJ, KW, MBM; Study Design and Protocol: KW, BT, CT, JJ, WW; Study Protocol revisions: all authors; Ethics/Governance: KW, AL; Site chief investigators: HA, GB, PB, KW, AS; Data collection: AL, HA, PB, KW, AS; Data analysis: JJ, CT, BT; Manuscript: KW, JJ, CT, BT; Manuscript revisions: all authors; Manuscript guaranteed by KW and BT.

### Collaborators

Rachel Rosler: network sponsor

Melanie Stephenson: literature review

Kim Hansen: risk advisor

Ms Ella Martini: consumer

Dr Hamish Rodda: emergency informatics advisor, project sponsor

Dr Judy Lowthian: district nursing researcher

## Acknowledgements

Lisa Kuhn, Anne Spence, Cathie Piggot: governance assistance John Papatheohari, David Rankin: project sponsors and advisors Mrs Katarina Tomka: project facilitator, Monash University

