## Appendix for "Emergency medicine patient wait time multivariable prediction models: a multicentre derivation and validation study"

Appendix 1a. Collected and derived predictor variables.

| Collected predictor variables | Type of Variable |
|---|---|
| **Required to calculate triage-to-provider time** | |
| Triage time | Date/Time |
| First seen by provider time | Date/Time |
| **Variables required to calculate proposed predictor variables** | |
| Ambulance at destination/door (Front door time only available for ambulance patients) | Date/Time |
| Ambulance handover complete (Off stretcher time, only for ambulance patients) | Date/Time |
| Clinical decision to admit time (previous patients) | Date/Time |
| Date of Birth | Date/Time |
| Departure time (previous patients) | Date/Time |
| **Other predictor variables available for model development** | |
| Advanced care directive alert | Categorical |
| Arrival transport mode | Categorical |
| Campus code | Categorical |
| Compensable status | Categorical |
| Country of birth | Categorical |
| Indigenous status | Categorical |
| Interpreter required | Categorical |
| Preferred language | Categorical |
| Referred by | Categorical |
| Gender | Categorical |
| Triage category (Australasian Triage Scale) | Categorical |
| Type of usual accommodation | Categorical |
| Type of visit | Categorical |
| **Calculated predictor variables** | |
| Age (We used age rather than date of birth to preserve privacy) | Continuous |
| Patients in triage queue: The number of patients who arrived before the patient of interest but have not been triaged; requires Arrival and Triage date/time | Continuous |
| Patients awaiting a provider: The number of patients who have completed triage before the patient of interest, but have not yet seen a provider; requires Triage and First seen by provider Date/Time | Continuous |
| Admitted patients awaiting departure: The number of patients who have had an admission decision made and have not yet departed from the Emergency Department; requires Clinical decision to admit and departure date/time | Continuous |

| | |
|---|---|
| Ambulance offload queue: The number of ambulance patients arrived, but not yet off their stretcher; requires Ambulance at destination and Ambulance off-stretcher date/time | Continuous |
| Average wait-time of the last k-patients: The average calculation of the triage-to-provider time of the last k-patients that are seen by the provider prior to the patient of interest arriving; requires Triage-to-provider time of previous patients and First seen by provider of previous patients date/time plus the Triage date/time of the patient of interest | Continuous |

Appendix 1b. Incomplete data columns and number of episodes impacted

| Predictor variable | Descriptor | Number of episodes |
|---|---|---|
| Type of usual accommodation | Unknown/unable to determine | 613,474 |
| Arrival transport mode | Other | 1,349,317 |
| Compensable status | Compensable status unknown | 478,415 |
| Country of birth | Not Stated | 772,685 |
| Interpreter required | Not Stated / Inadequately Described | 2,942 |
| Referred by | Other | 232,848 |
| Preferred language | Not Stated | 1,040,001 |
| Indigenous status | Question unable to asked | 15,926 |

Appendix 1c. Full models: Internal validation of each site-specific, full model using its own hospital 2019 testing data; distributions of absolute errors for wait time predictions

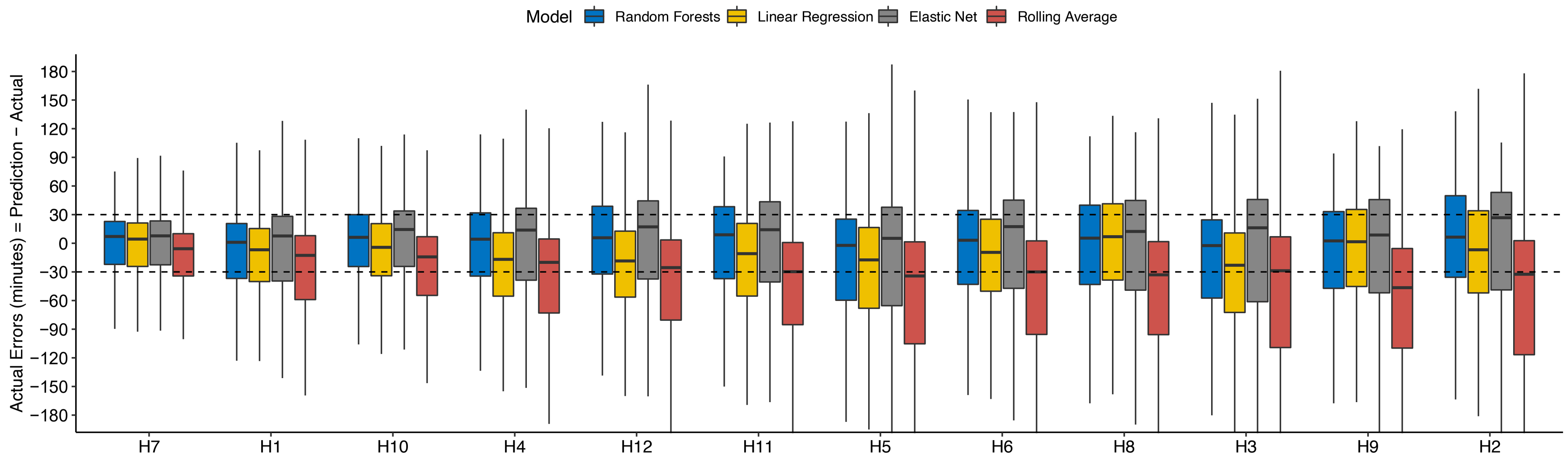

Box and whisker plots of median absolute errors of the wait−time estimates. The horizontal bold black line within each boxplot indicates the median value of the absolute errors. The box describes the interquartile range (IQR, Q1 and Q3). The lower whisker extends from Q1 to the lowest value at most Q1 − 1.5xIQR. The upper whisker extends from Q3 to the highest value at most Q3 + 1.5xIQR. The horizontal dashed line indicates the acceptable threshold of +/− 30 minute wait time estimate errors. Each box colour represents a different model generation technique.

Appendix 1d. Simplified models: Internal validation of each site-specific, simplified model using its own hospital 2019 testing data; distributions of absolute errors for wait time predictions

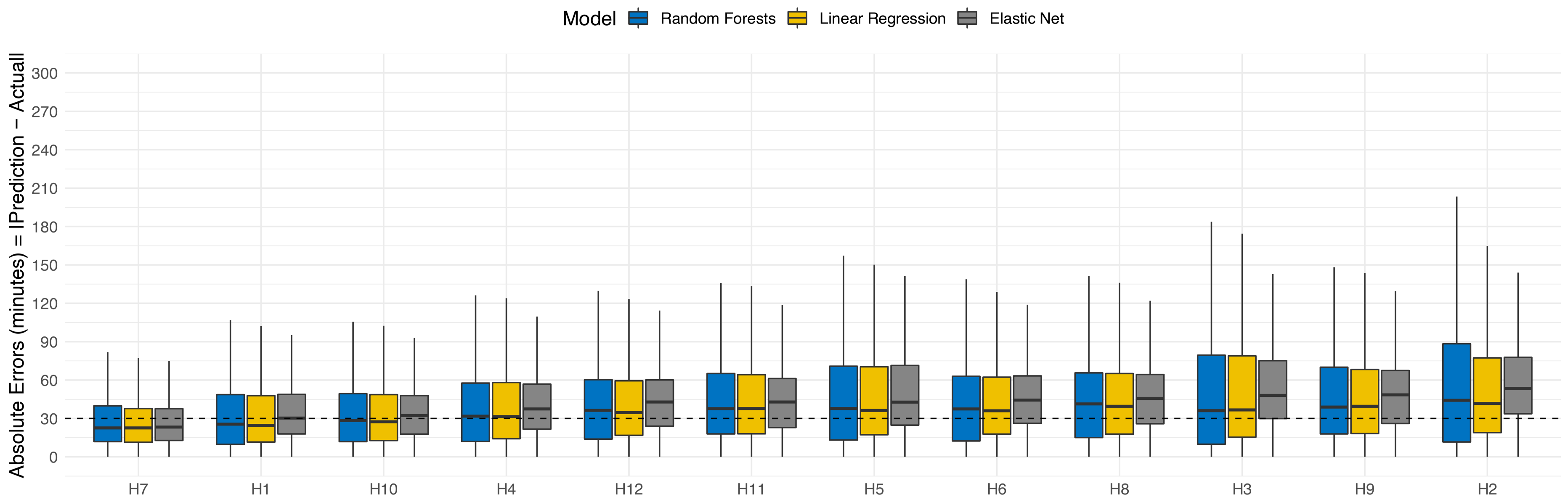

Simplified models built with top−ranked variables that account for 95% of the relative importance.

Appendix 1e. Cross-site, site-specific comparisons: distributions of absolute errors

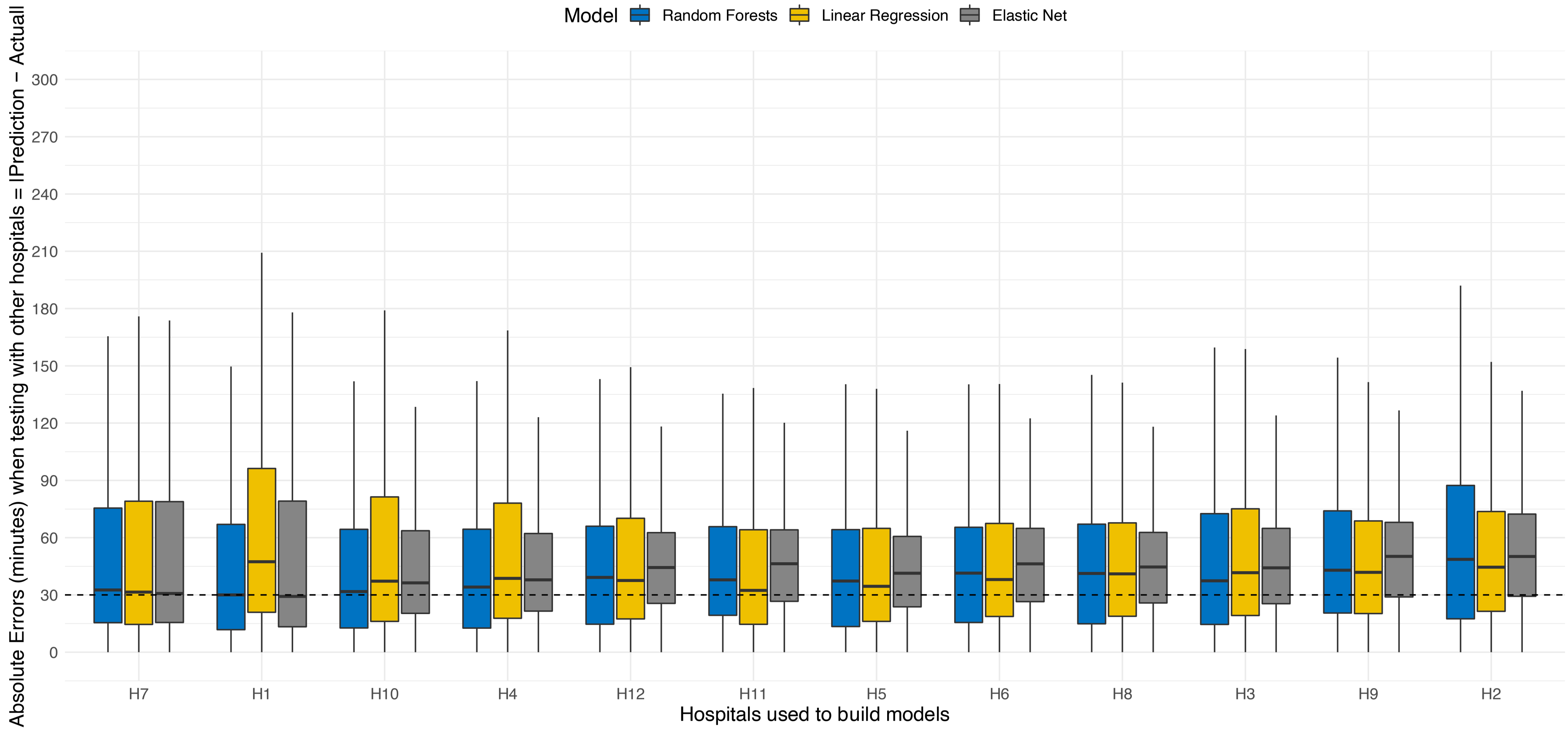

Appendix 1f. Distributions of absolute errors for wait time predictions before and during COVID19 reduced attendances in 2020

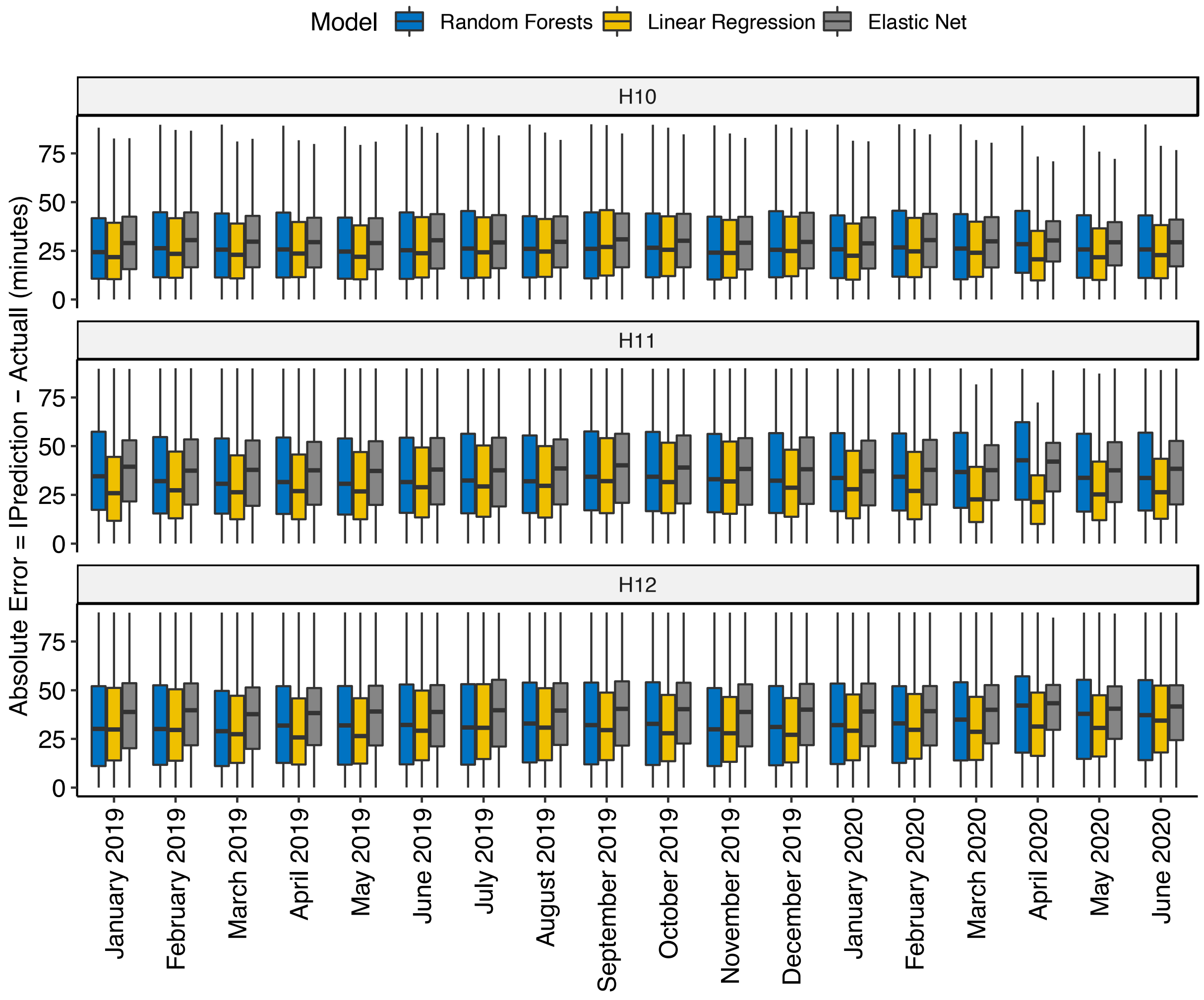

| ED presentations over time | H10 | H11 | H12 |
|---|---|---|---|
| January 2019 | 3164 | 5598 | 4478 |
| February 2019 | 3104 | 5109 | 4173 |
| March 2019 | 3346 | 5903 | 4771 |
| April 2019 | 3325 | 5440 | 4613 |
| May 2019 | 3449 | 5865 | 4836 |
| June 2019 | 3310 | 5592 | 4658 |
| July 2019 | 3323 | 5643 | 4722 |
| August 2019 | 3455 | 5832 | 4675 |
| September 2019 | 3332 | 5509 | 4440 |
| October 2019 | 3367 | 5685 | 4736 |
| November 2019 | 3312 | 5405 | 4621 |
| December 2019 | 3437 | 5546 | 4663 |
| January 2020 | 3259 | 5665 | 4565 |
| February 2020 | 3252 | 5261 | 4607 |
| March 2020 | 3063 | 7369 | 4107 |
| April 2020 | 2200 | 5538 | 3177 |
| May 2020 | 2460 | 4425 | 3874 |
| June 2020 | 2714 | 4846 | 3932 |

*The first wave of COVID−19 in Victoria occured from March 2020
